# Anthropozoonotic spillovers reveal sustained long-term cryptic circulation of SARS-CoV-2 within and between Lithuanian mink farms

**DOI:** 10.1101/2025.07.15.25331253

**Authors:** Martynas Smičius, Ingrida Olendraitė, Jonas Bačelis, Aistis Šimaitis, Miglė Gabrielaitė, Bas B Oude Munnink, Reina S Sikkema, Arūnas Stankevičius, Žygimantas Janeliūnas, Paulius Bušauskas, Egidijus Pumputis, Simona Pilevičienė, Petras Mačiulskis, Marius Masiulis, Vidmantas Paulauskas, Snieguolė Ščeponavičienė, Monika Katėnaitė, Rimvydas Norvilas, Ligita Raugienė, Rimvydas Jonikas, Inga Nasvytienė, Živilė Žemeckienė, Kamilė Tamušauskaitė, Milda Norkienė, Emilija Vasiliūnaitė, Danguolė Žiogienė, Albertas Timinskas, Marius Šukys, Mantas Šarauskas, Dovilė Juozapaitė, Daniel Naumovas, Arnoldas Pautienius, Astra Vitkauskienė, Rasa Ugenskienė, Alma Gedvilaitė, Darius Čereškevičius, Laimonas Griškevičius, Marion Koopmans, Alvydas Malakauskas, Gytis Dudas

## Abstract

Several studies have documented reverse zoonotic transmission of SARS-CoV-2, including in farmed mink which are susceptible to human respiratory viruses and are known for serving as a reservoir capable of generating new virus variants in densely populated farms. Here, we present the results of a genomic investigation launched in response to detected human infections with mink-origin SARS-CoV-2 lineages, and show evidence of at least 14 high-confidence introductions of SARS-CoV-2 from humans into farmed mink in Lithuania where sustained transmission in farmed mink lasted up to a year. We estimated the most likely timeframes for these introductions encompassing at least six SARS-CoV-2 lineages, some of which were already extinct in humans, with Bayesian phylogenetic and molecular clock analyses. This study highlights the public health risks posed by fur farms and underscores that passive genomic surveillance systems are ineffective without the active involvement and expertise of responsible institutions.

## Introduction

### SARS-CoV-2 spreads in mink farms

SARS-CoV-2 is a zoonotic betacoronavirus that emerged in humans in China in late 2019, causing the COVID-19 pandemic and continues to circulate in humans and other vertebrate species endemically - both wild and domestic (V’kovski et al., 2021). Mink are generally susceptible to respiratory viruses, including human or avian influenza viruses (Sun et al., 2021) while also being farmed industrially in dense populations for their furs. Quite early in the COVID-19 pandemic, SARS-CoV-2 was detected in mink farms (Netherlands) and later in a plethora of countries (including Spain, Denmark, the United States, Sweden, Italy, France, Greece, Lithuania, Canada, Poland, Latvia) (Jahid et al., 2024; Oreshkova et al., 2020). In addition to human-to-mink spillovers, anthropozoonotic spillovers from animals to humans were also detected and documented, as seen in the Netherlands and Denmark, including transmission between farms (Larsen et al., 2021; Oude Munnink et al., 2021). Governments responded in different ways: some, like Denmark and the Netherlands performed mass culling of entire mink farms due to concerns of spillover back into the human population, while others, including Canada and Lithuania, tried to contain the transmissions through testing, isolation and disinfection (Jahid et al., 2024).

### Mink farms in Lithuania

In Lithuania, farmed mink products have accounted for between 20.9 and 75.3 million euros worth of exports yearly, depending on the year (2012–2022) (“2023/0376/LT (Lithuania),” n.d.). The country is among the 10 largest fur producers globally, producing more than a million pelts per year between 2019–2022, decreasing to 600,000 pelts in 2023 (“Fifur Statictics,” 2025). Biosecurity requirements issued by the State Food and Veterinary Service of the Republic of Lithuania (SFVS), the main government body responsible for food and veterinary safety in Lithuania, have been compulsory on mink farms in the country since 2015 (“B1-432,” n.d.) and include fencing, pest control, hygiene stations (see Supplementary materials section “Biosecurity requirements in Lithuanian mink farms”).

During the SARS-CoV-2 pandemic, despite evidence of at least three officially recognised outbreaks on mink farms in late 2020, no mass culling was performed, and the standard operating procedures of the SFVS regarding SARS-CoV-2 in animals lacked strict conditions and definitions to enable decisive action. Regulations included statements requiring “good zoonotic disease status” in affected farms but with no mandatory periodic testing of farms to detect infections and self-reporting of increased mortality or morbidity (“B1-850,” n.d.; “B1-991,” n.d.; Žigaitė et al., 2023, pp. 2020–2021). Upon detection of any disease in a farm, an SFVS permit is required for any movement of animals between farms (“I-2110 Lietuvos Respublikos veterinarijos įstatymas,” n.d.). As of December 2021, before planned animal transfer, the SFVS has to be informed about the “zoonotic disease status” in the farm. The SFVS also has to be informed about any cases of confirmed or suspected SARS-CoV-2 infections among farm employees, prior to planned animal transfer. However, there is no mandatory testing of animals for SARS-CoV-2 before the transfer if a farm’s “zoonotic disease status” is considered acceptable.

### Evolutionary changes in animal RNA viruses

*De novo* evolution towards increased pathogenicity is well described for avian influenza viruses in domesticated hosts kept at high densities, e.g. H5 and H7 viruses have been shown to quickly adapt to infecting poultry (Banks et al., 2001; Monne et al., 2014). Avian influenza viruses predominantly circulate as low pathogenicity variants in wild birds, with high pathogenicity forms evolving readily after spillovers into domestic birds via acquisition of polybasic cleavage sites in their hemagglutinins (Dhingra et al., 2018; Luczo et al., 2015). Once avian influenza becomes highly infectious, it spreads rapidly and widely through the poultry population, leading to a high risk of spillover to mammals due to increased exposure (Plaza et al., 2024). Adaptations to these new mammalian hosts happen readily and are well-described (Kim et al., 2025). Similar increases in pathogenicity are observed in other RNA viruses. Infectious salmon anaemia (ISA) virus is an orthomyxovirus that was first detected in farmed salmon, another domesticated species grown in high density conditions, and can readily evolve into high pathogenicity forms through deletions that affect the length of their surface protein haemagglutinin esterase (HE) stalk, leading to both low and high pathogenicity forms co-circulating in the same populations (Godoy et al., 2013; Plarre et al., 2012; Rimstad and Markussen, 2020).

While several animal species are susceptible to SARS-CoV-2, farmed mink stand out due to their dense and open housing conditions, high population and contact with humans. Escape of such variants from farms through infected employees or environmental contamination (Chaintoutis et al., 2021) can lead to increased risk of unpredictable outcomes to human public health.

For example, some studies suggest that the B.1.1.529/Omicron variant might have evolved in rodents, due to some of the mutations carried by this lineage increasing Spike’s affinity to mouse ACE2 receptors (Cameroni et al., 2022; Wei et al., 2021). On the other hand, there is evidence that mink-adaptive mutations in SARS-CoV-2’s Spike protein does not increase its fitness in humans (Zhou et al., 2022).

With continued circulation of SARS-CoV-2 in farmed mink, a new mink-associated virus may eventually spill back into human populations. In Denmark, a single clade of lineage B.1.1.298 derived from mink was responsible for a substantial number of human cases in the region surrounding a mink farm, illustrating the potential of the virus to escape and infect humans (Fenollar et al., 2021; Rasmussen et al., 2024). Additionally, studies have shown that these farm-origin viruses can potentially spill over into local wildlife, further highlighting their escape potential (Strang et al., 2022). Moreover, there is evidence that outbreaks in farmed mink populations can be sustained for at least 14 months while staying undetected (Domańska-Blicharz et al., 2023).

### SARS-CoV-2 mink and human surveillance in Lithuania

In November 2020, after SARS-CoV-2 was detected in Danish mink farms, the SFVS started a passive surveillance program for mink farms in Lithuania.The program was based on mandatory reporting by farmers of increased mink mortality or morbidity with symptoms like fever, reduction in feed intake, any signs of respiratory or digestive disorder, together with any confirmed cases of SARS-CoV-2 infection among farm employees (Žigaitė et al., 2023). In addition, farms had to report live, diseased, and dead mink numbers weekly, while SFVS strengthened supervision of existing biosecurity measures on farms, and introduced a requirement for farm staff to wear personal protective equipment while working with mink. The first two farms with SARS-CoV-2 infection among mink were detected in November and December 2020, soon after the beginning of passive surveillance (Žigaitė et al., 2023, pp. 2020–2021) with additional two infected mink farms detected at the beginning of 2021 (Žigaitė et al., 2023, pp. 2020–2021). Instead of culling affected farms, the SFVS created and approved an outbreak control and mink culling plan for infected farms consisting of culling all SARS-CoV-2-positive or possibly infected mink as well as mink in cages surrounding them; disinfection of such cages and their surrounding area, compulsory disinfection of vehicles leaving the farm, improved surveillance of farm staff health status, and increased use of PPE.

### SARS-CoV-2 lineage composition in mink and humans in Lithuania

Human SARS-CoV-2 surveillance in Lithuania before March 2021 was limited in scope and relied on opportunistic sampling and sequencing, with the first sequences from the country appearing on GISAID in October 2020 (Khare et al., 2021). Routine and representative surveillance in Lithuania was launched in March 2021. Available data indicate that from October 2020 to February 2021, i.e. prior to routine surveillance, B.1.177.60 and other B lineages (notably B.1.1.280) were the most prevalent in humans in Lithuania. Later, in March 2021, these lineages were pushed out by Alpha lineages (B.1.1.7, its sublineage Q.1 and their relatives). Alpha-like lineages stayed dominant until July 2021, when case numbers receded during the summer. Afterwards, Delta/B.1.617.2 drove a wave of COVID-19 cases that established these lineages as the predominant SARS-CoV-2 variant in the country (Delta-descendant AY.4.5 lineage in particular). Delta-like lineages stayed dominant until January 2022, when Omicron/B.1.1.529 variants swept the world, pushing other SARS-CoV-2 lineages previously circulating in humans to apparent extinction.

On 2021 October 05, at a time when the Delta/B.1.617.2 lineage was dominant globally (including in Lithuania), routine human SARS-CoV-2 surveillance discovered a mink farm worker infected with lineage B.1.343, which hadn’t been seen in humans in Lithuania since December 2020. A similar case of B.1.177.60, an extinct lineage of Lithuanian origin with mink-adaptive mutations from a farm worker, was detected a few weeks later (2021 November 11). Upon communication of this situation by representatives of the Lithuanian genomic surveillance programme to the Lithuanian government and SFVS, a recommended country-wide testing of mink farms was carried out in 2021 October–November. All 57 mink farms in the country were tested without prior warning and SARS-CoV-2 infections were detected in 25 farms, with RT-PCR (13) or ELISA (16) (Žigaitė et al., 2023).

Here we describe an investigation that combines sequence data derived from human SARS-CoV-2 surveillance efforts and sequencing of SARS-CoV-2 from affected mink farms after the country-wide test, highlighting the need and benefits of active pathogen surveillance through sequencing in vertebrate hosts known to be susceptible to SARS-CoV-2 and other pathogens.

## Methods

### SARS-CoV-2 genomic surveillance programme

Briefly, the genomic surveillance programme of Lithuania combined the sequencing capacity of Vilnius University Life Sciences Center, Vilnius University Hospital Santaros Klinikos, Lithuanian University of Health Sciences, Lithuanian University of Health Sciences Hospital Kauno Klinikos, and the European Center for Disease Control (ECDC). For details about each institution’s sequencing infrastructure and bioinformatic processing see (Dudas et al., 2021). This SARS-CoV-2 genomic surveillance programme averaged over 600 genomes per week (translating to the ability to exclude the circulation of a lineage at a weekly 0.005 frequency with 0.95 probability (Brito et al., 2022)). From the inception of this programme in March 2021, by sampling design, all SARS-CoV-2 PCR-positive samples are directed to sequencing if a person is registered as working on a fur farm. The SFVS had no specific genomic surveillance design for mink and sequencing was only done in response to confirmed outbreaks in mink farms. The number of SARS-CoV-2 cases was obtained from the official state statistics portal (https://osp.stat.gov.lt/covid-dashboards), administered by the National Data Agency of Lithuania (“ZUDC,” n.d.).

#### Mink-origin SARS-CoV-2 genomes

SARS-CoV-2 samples from mink were collected by the SFVS and sequenced by the National Food and Veterinary Service of Lithuania or Erasmus University Medical Center. These were mapped to a reference SARS-CoV-2 genome using Dragen (Version 4.3, Illumina) and submitted to GISAID (Khare et al., 2021). Upon inspection, the quality of these genomes was found to be insufficient, and they were remapped from raw FASTQ files using COVID-19-SIGNAL pipeline (Nasir et al., 2024) under default parameters. A total of 58 mink samples were used for this study, with Pangolin (O’Toole et al., 2021) assigning the 58 genomes to 6 broader lineages (see next section): 30 to AY.4, 4 to AY.122, 3 to B.1.1.7, 9 to B.1.1.280, 5 to B.1.177.60, and 7 to B.1.343 (Supplementary material file “Sample_table_with_EPI_ISL.csv”).

Data about mink farms and herd sizes was sourced from the Agriculture Data Center website (https://zudc.lt/ukiniu_gyvunu_registras/menesio-ataskaitos/) (“COVID-19 Lietuvoje,” n.d.).

### Contextual data

Contextual data for molecular clock analyses were generated on a lineage-by-lineage basis (split into B.1.1.7, AY.4, AY.122, B.1.177.60, B.1.1.280, and B.1.343 categories) by uploading the reassembled mink-origin SARS-CoV-2 genomes of each lineage to UShER (Turakhia et al., 2021) and noting the genomes most closely related to these focal sequences which we called the “mink-adjacent” sequences. For broader context, a metadata file was downloaded from GISAID for all sequences on the database (Shu and McCauley, 2017). For each sequence in the database, we assigned one of our six focal lineage categories, i.e. if a sequence was assigned to lineage Q.1 or Q.2 (i.e., B.1.1.7.1 or B.1.1.7.2) it was considered to be in the B.1.1.7 category. Due to occasional lineage misassignments on GISAID, every sequence belonging to a given lineage had to fall within a known circulation window of that lineage (B.1.1.7: 2020 Oct–2021 Oct, AY.4: 2021 May–2022 Feb, AY.122: 2021 May–2022 Feb, B.1.177.60: 2020 Oct–2021 May, B.1.1.280: 2020 Aug–2021 Mar, B.1.343: 2020 Mar–2020 Sep). Due to different lineage prevalences (globally and locally), we chose different numbers to sample at random from each lineage category (1 sequence per country per month for B.1.1.7, AY.4 and AY.122, 5 sequences per country per month for B.1.177.60, and 10 sequences per country per month for B.1.1.280 and B.1.343). For some lineages, 1 sequence per country per month resulted in datasets that were too large to analyse in BEAST, and in those cases, we randomly downsampled, by randomly removing contextual genomes from the dataset, reducing them to 40% (B.1.1.7), 50% (AY.122), or 60% (AY.4) of their original size. Due to low numbers of B.1.1.280 and B.1.343 lineage genomes overall, additional B.1.1 and B.1 sequences (respective ancestral lineages), respectively, were added to each category from GISAID, with 25 B.1.1 sequences per month added to the B.1.1.280 category and 35 B.1 sequences per month added to the B.1.343 category. After contextual sequence generation, the sequences were aligned using Nextclade version 3.8.2 (Aksamentov et al., 2021) to reference genome NC_045512.2.

### Phylogenetic Analysis

Each of the six lineage datasets (B.1.1.7 (256 genomes), AY.4 (266 genomes), AY.122 (224 genomes), B.1.177.60 (247 genomes), B.1.1.280 (178 genomes), and B.1.343 (210 genomes)) was used for analysis with BEAST v1.10.4. XML files for these analyses were generated using BEAUti v1.10.4. with the following parameters: 1) SRD06 substitution model (Shapiro et al., 2006) that models substitutions separately depending on codon position (one model for the first two codon positions and another for the third position) according to two independent Hasegawa-Kishino-Yano (HKY) + Γ_4_ substitution models (Hasegawa et al., 1985; Yang, 1994) with gene coordinates provided in the XML. The HKY model assumes different substitution rates for transitions and transversions and unequal nucleotide frequencies, while Γ assumes rate heterogeneity across sites with a gamma distribution discretised into 4 rate categories; 2) an uncorrelated relaxed clock with log-normally distributed rates calibrated on tip dates (Drummond et al., 2006) and a continuous time Markov Chain reference prior (Ferreira and Suchard, 2008) on the molecular clock rate. A relaxed clock allows substitution rates to vary among branches in the tree, and in uncorrelated clock models the substitution rates are not correlated across neighboring branches. Each sequence is associated with a specific sampling date, therefore, the molecular clock is tip-calibrated; 3) Coalescent Bayesian SkyGrid tree prior (Gill et al., 2013) that allows for a varying effective population size across different time periods. The default gamma prior with shape 0.001 and scale of 1000 was used, favoring low population sizes with high variability. The SkyGrid number of grid points was set to 9 time points and the cut off point to 1.5 years prior to the most recent tip date; 4) 200 million steps of MCMC with sampling every 20,000 steps. For each lineage, three replicate chains were run and convergence assessed using Tracer v.1.7.2 (Rambaut et al., 2018). Convergence was assessed based on all parameters having effective sample sizes (ESSs) above 200; 5) Host information was included as a discrete trait and reconstructed using an asymmetric substitution model inferred by Bayesian stochastic search variable selection (BSSVS) (Gill et al., 2013). To obtain Markov jumps (transitions between human and mink as a host), we used complete history logging (Gong et al., 2013; Minin and Suchard, 2008a), generating an MCMC output file with history parameters for nodes and leaves, containing Markov jump dates. 6) As we wanted to estimate the number of SARS-CoV-2 introductions into mink, we set the host trait of five human-origin samples as mink to assist ancestral state reconstruction: S21L465|Lithuania (EPI_ISL_7083492), S21L477|Lithuania (EPI_ISL_7082794), S21E887|Lithuania (EPI_ISL_2428956), S21E881|Lithuania (EPI_ISL_2428882), IBT-LCS-VU_r24_28|Lithuania (EPI_ISL_5390697). These samples are exceedingly likely to come from mink-to-human spillovers as they contain mink adaptive mutations and were not detected in the general human population at the time yet strongly and incorrectly inform ancestral state reconstruction due to lack of samples from mink. This is discussed in greater detail in results. 7) Sequences in the XML file were modified by inserting two ‘N’ nucleotide symbols after position 13 468 to bring ORF1b into frame with ORF1a.

### Post-processing of MCMC samples

For each BEAST analysis, three replicate chains were run to ensure convergence to the posterior distribution. The one exception were analyses of lineage B.1.1.280, where the posterior distribution appeared to be bimodal for parameters related to the evolutionary rate - age(root), treeLength, default.ucld.mean, default.meanRate and skygrid. We ran B.1.1.280 XMLs an additional eight times with the same bimodal behaviour. Due to this bimodal behavior, we decided to use only 6 of the 11 runs (with age(root) around 2020.07 and treeLenth around 47.9 years), with a burn-in of 75% (150 million states) to generate a workable dataset. All log and tree files are provided in supplementary materials.

### Calculation of human to mink transmission times

For each lineage, MCMC output trees were combined with LogCombiner(v1.10.4) with burn-in removed, corresponding to 10% of the generated output trees or 20 million initial trees, leaving 27 000 trees for analysis in each case (75% or 150 million initial trees for B.1.1.280, leaving 15 000 trees for analysis). From these combined trees, maximum credibility clade trees were generated with TreeAnnotator(v1.10.4). For each human-to-mink transmission event which was visually identified from the tree, the most recent common ancestor (MRCA) was identified together with all the descendant leaves. For further analysis, MCMC output trees were used, by taking only the MRCA nodes with leaves identical to leaves in the MCC (maximum clade credibility) tree, i.e. conditioning our analysis on the clades present in the MCC tree that represent SARS-CoV-2 introductions into mink. From these clades, introduction times from humans into mink were extracted and summarised.

### Visualisation and Filtering

All visualizations were created using Python (3.12.8) and several packages (matplotlib 3.10.1, cartopy 0.24.0, shapely 2.1.0, baltic 0.3.0). SNP (single nucleotide polymorphism) alignments with phylogenetic trees were generated by taking a branch from the MCC tree for each lineage which includes the leaves of interest regarding human-to-mink transitions. The first and last 50 nucleotides, synonymous mutations and identical nucleotides were omitted.

### Case numbers and lineage proportions

Case number and lineage proportions were visualised with matplotlib (version 3.9.2) in Python. For a period from March to May and September to November in 2021, more than 6 million (6 376 146) antigen and PCR tests were performed in Lithuania, with a test positivity rate of 8.3% (527 082), and a rate of 3.3% (17 345) for sequencing. This time period covers the beginning of genomic surveillance in Lithuania in early 2021, capturing the tail-end of the winter wave and arrival of Alpha-like lineages (March to May), skips the summer with low COVID-19 case numbers and extremely high sequencing coverage, and resumes in late 2021 (September to November), encompassing the wave driven by Delta-like lineages and before the arrival of Omicron-like lineages. Given these acceptable surveillance parameters, we assumed that lineage proportions of SARS-CoV-2 genomes from Lithuania on GISAID were representative; as such, we scaled these lineage proportions to total SARS-CoV-2 case numbers in Lithuania. The lineage proportions and case numbers were aggregated into 14-day periods.

### Estimating the number and dates of human-to-mink transmissions

For each focal lineage analysis after discarding 20 million states as burn-in (with additional processing for lineage B.1.1.280, outlined above), we extracted the human-to-mink Markov jump (Minin and Suchard, 2008b) dates from each tree sampled during MCMC. Since many of the human-to-mink transition dates were multimodal, we used an algorithm that uses a kernel density estimate (KDE) of the jump dates and simulates a vertically descending line that intersects the KDE at multiple points and computes the integral until it reaches 0.95 (i.e. the 95% highest posterior density interval). Following the same burn-in processing, we also extracted the number of human-to-mink Markov jump state changes from the posterior.

## Results

We analysed 1 323 SARS-CoV-2 genomes from humans and 58 from mink to assess the effectiveness of Lithuania’s genomic surveillance programme and to evaluate the risks associated with mink farming. We identified mink-associated human infections bearing mutations associated with mink adaptation, commonly affecting mink farm employees and caused by SARS-CoV-2 lineages that have been extinct in the general human population for months.

### Correcting ancestral state reconstruction

During initial runs, we noticed that three lineage B.1.343 spillback events into humans were strongly and incorrectly informing ancestral state reconstruction at neighbouring nodes (i.e., inferring internal nodes of the mink-associated subtree as human). There is abundant evidence that this is incorrect: 1) the last time lineage B.1.343 was seen in the human population of Lithuania was in December 2020 (EPI_ISL_934083), while its reemergence in May 2021 (EPI_ISL_2428882) and October 2021 (EPI_ISL_5390697) happens at a time when major global variants of concern (VOCs) Alpha and Delta had outcompeted virtually all co-circulating SARS-CoV-2 variants; 2) two of the samples (EPI_ISL_5390697, EPI_ISL_2428882) contain the ORF3a:L219V mutation, and all three samples contain S:F486L and ORF1a:G4177E mutations associated with mink adaptation, and 3) none of these mutations are detected in an earlier human sample from December 2020 (EPI_ISL_934083).

A similar situation was noticed for lineage B.1.177.60, where the last human infections with this lineage in Lithuania were detected in June 2021 (EPI_ISL_3060127) and subsequent detections were in mink farmers or mink in November 2021, at a time when Delta variants were dominant. Additionally, the ORF3a:L219V mutation is present in both human and mink samples from November 2021. At the time when these suspected zoonotic spillover events were detected in the Lithuanian population, there were no reported sequences of lineages B.1.343 and B.1.177.60 anywhere in the world (Figure 1).

**Figure 1.**
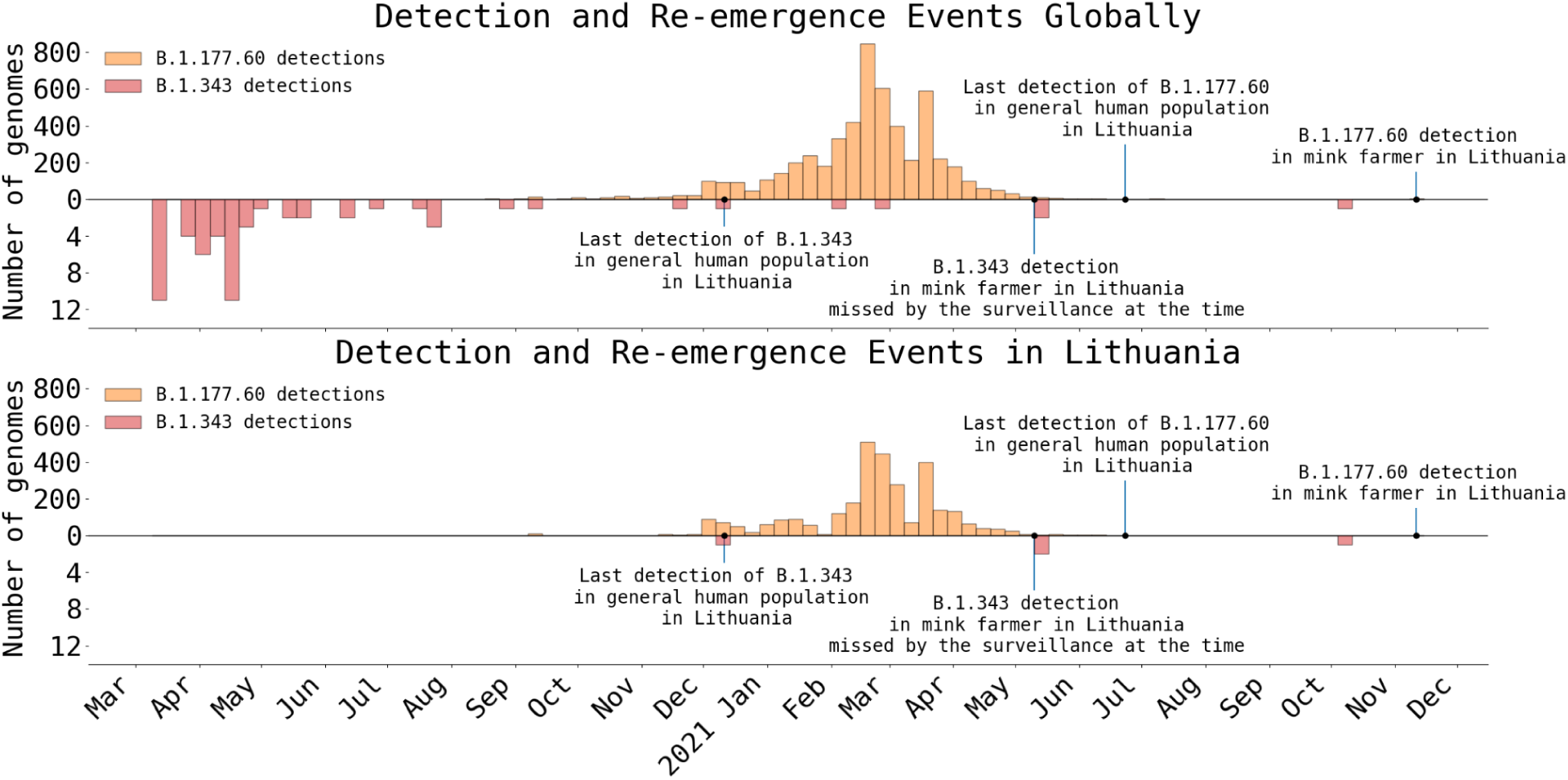
The number of B.1.343 and B.1.177.60 lineage genomes globally (top) and in Lithuania (bottom), based on data from GISAID. Genomes from human hosts are shown. Bars correspond to weekly counts.

In both cases, continued circulation of SARS-CoV-2 in mink rather than humans is more parsimonious. For lineage B.1.343, the Lithuanian SARS-CoV-2 genomic surveillance programme sequenced 26 258 SARS-CoV-2 genomes out of 802 052 human cases between 2020 December 11 (last known human B.1.343 infection, EPI_ISL_934083) and 2021 November 30 (first putative spillback from mink into humans, EPI_ISL_10571268) which corresponds to a maximum probable prevalence of 9.57×10^-5^ in the absence of lineage detection (Brito et al., 2022). For SARS-CoV-2 to have sustained itself over this nearly year-long period exclusively via human-to-human transmission and without detection, B.1.343 could have caused, at most, 2.5 human cases which is highly unrealistic. For lineage B.1.177.60, the equivalent calculation is: last detected human case 2021 June 23 (EPI_ISL_3060127), first putative spillback 2021 November 11 (EPI_ISL_3060127) with 9419 SARS-CoV-2 genomes sequenced from 347 942 human cases over this time without detections and therefore a maximum probable prevalence of 2.67×10^-4^ in the human population. For sustained and undetected human-to-human transmission this would require around 93 human cases over 141 days which is also questionable.

Due to the reasons outlined above, the host trait for sequences (S21L465|Lithuania (EPI_ISL_7083492), S21L477|Lithuania (EPI_ISL_7082794), S21E887|Lithuania (EPI_ISL_2428956), S21E881|Lithuania (EPI_ISL_2428882), IBT-LCS-VU_r24_28|Lithuania (EPI_ISL_5390697) was assigned as “mink” rather than “human” while preparing XML files for BEAST analysis.

### Mink population dynamics in Lithuanian farms

The requirements and regulations for mink farms in Lithuania were ambiguous and not strictly regulated. As such, neither the farmed mink nor the spread of SARS-CoV-2 during the COVID-19 pandemic were markedly impacted by public health responses (Figure 2). Mink farming is seasonal with known breeding and pelting times, leading to regular population spikes. The decrease in mink population before 2020 may be associated with decreased pelt prices. Likewise, a subsequent increase in population prior to 2021 could be associated with increased pelt prices, potentially affected by mink culling in Denmark and the Netherlands, i.e. decreased global pelt supply. Gradual annual decrease of the mink population since 2021 can be attributed to the ban of fur farms (first proposed in the parliament in November 2021 (n.d.)) in Lithuania that is supposed to take effect in 2027 (“VIII-500,” n.d.).

**Fig. 2.**
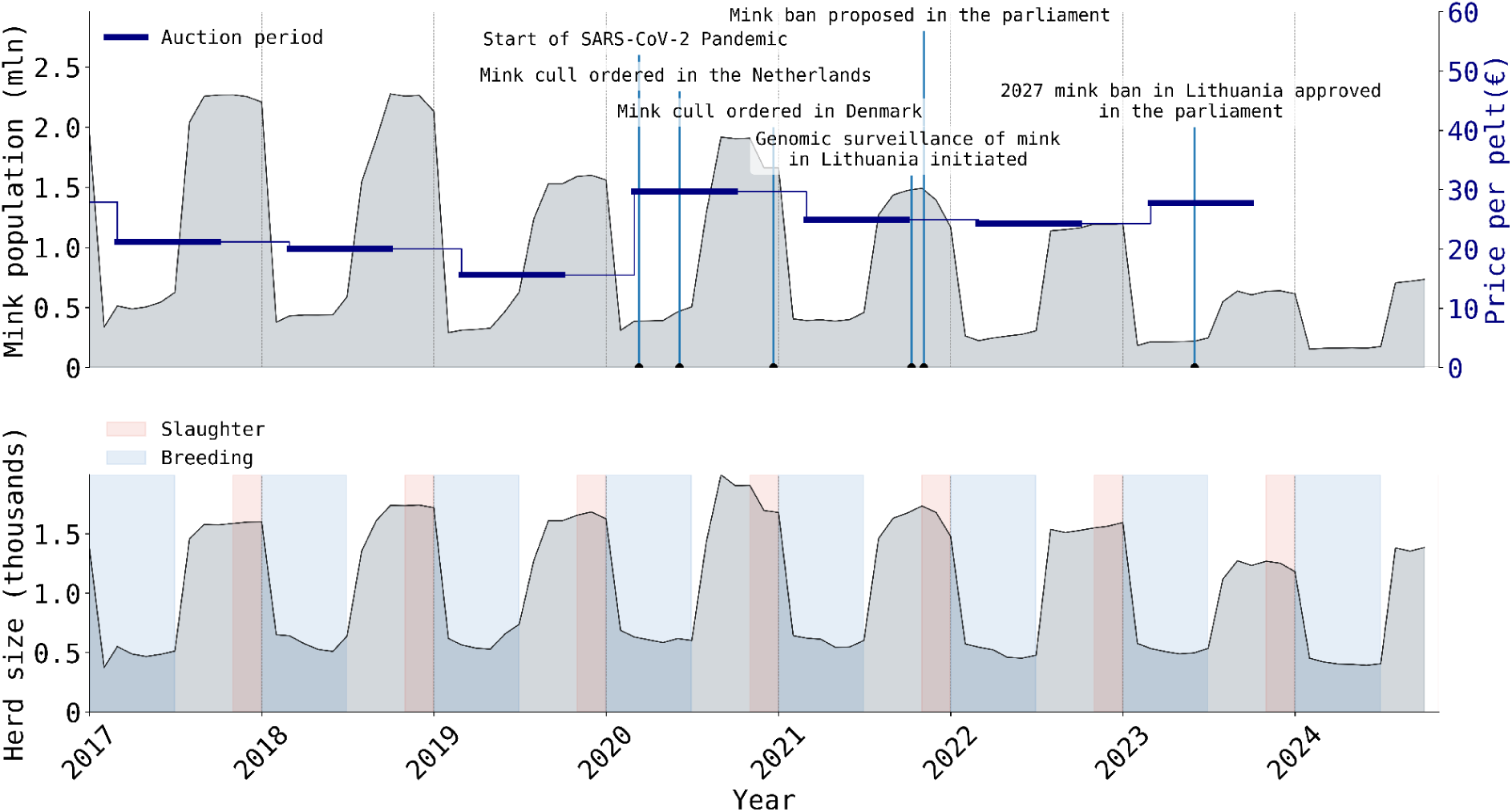
Mink population dynamics over time in Lithuania. The total mink population in Lithuanian mink farms with average yearly pelt price (auctions usually happen in March, June/July and September) on the regionally largest Saga Furs market (top) and the average mink herd size, as well as breeding with slaughter seasons in Lithuania from 2017 to 2024 (bottom). The number of animals is indicated on the Y-axis, and time is indicated on the X-axis. Data is registered monthly. Price data from FiFur (“Fifur Statictics,” 2025).

### Lineages transmitted from humans to mink

#### B.1.1.280 and B.1.343

The number of sequenced genomes of lineages B.1.1.280 and B.1.343 in Lithuania is limited as routine SARS-CoV-2 surveillance started only after these lineages went extinct in humans (Figure 3). B.1.1.280 was a largely Lithuania-restricted pre-Alpha lineage circulating in mid-to-late 2020 while B.1.343 was an obscure lineage mostly circulating in Denmark in 2020. These lineages were initially detected in Lithuania during sporadic sequencing efforts in September 2020 and December 2020 before routine surveillance started in March 2021 (Dudas et al., 2021). Mink-associated B.1.343 cases appeared in late 2021, both in mink and mink farmers but not in the general human population. Notably, there are two infected mink farm worker cases of B.1.343 in May 2021 that were not identified as mink-associated by the SARS-CoV-2 genomic surveillance programme in Lithuania at the time due to co-circulation of non-VOC lineages and only identified as such in October 2021, after the third spillback was discovered. By analysing posterior outputs from BEAST we infer that human-to-mink spillover for lineage B.1.343 happened once between October and December 2020 (95% HPD: 2020 Oct-09–2020 Dec-17; Figure 3, Figure 4). The mink farm worker cases discovered to be infected with lineage B.1.343 also contained some known adaptations to mink (ORF3a:L219V, S:F486L, ORF1a:G4177E) and were phylogenetically close to genomes from mink (Figure 5). Similarly, lineage B.1.1.280 also seems to have spilled over once, between June and September 2020 (95% HPD: 2020 June-26–2020 Sep-21; Figure 3, Figure 4) when the farmed mink population was high. For both of these lineages, the spillover events occurred before the start of representative surveillance, which should capture the trends in lineage distribution in the general human population in Lithuania.

**Figure 3.**
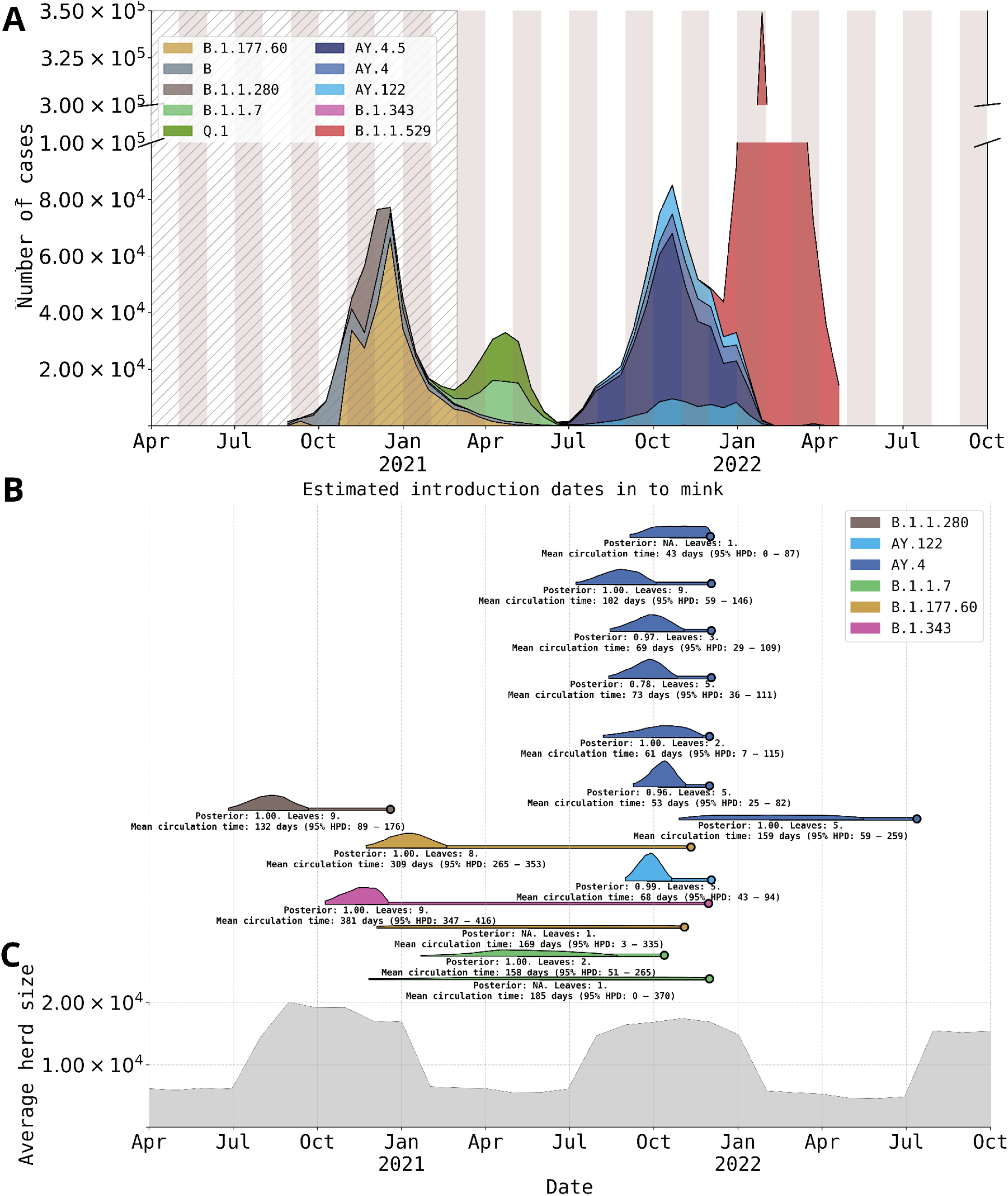
**A)** All reported COVID-19 cases in humans in Lithuania, scaled by lineage frequencies. Lineages are grouped into lineage groups (described in methods), the hatched area represents the time span prior to the establishment of routine SARS-CoV-2 genomic surveillance system in the country. **B)** The 95% highest posterior density (HPD) intervals for estimated dates of SARS-CoV-2 introductions into mink, conditioned on events detected in maximum credibility clade trees. The coloured line continuing after the 95% HPD signifies the continued circulation period until the last detected sample, which is marked by the dot at the end of the line. The text below each line marks the circulation period in days (starting date picked as the earliest in 95% HPD of estimated dates of SARS-CoV-2 introductions into mink and the days counted until the last detected sample) and the posterior probability for a given node (does not apply if there was only a single sample in mink for a transmission event). **C)** The average mink population per herd in Lithuania over the same time period.

**Figure 4.**
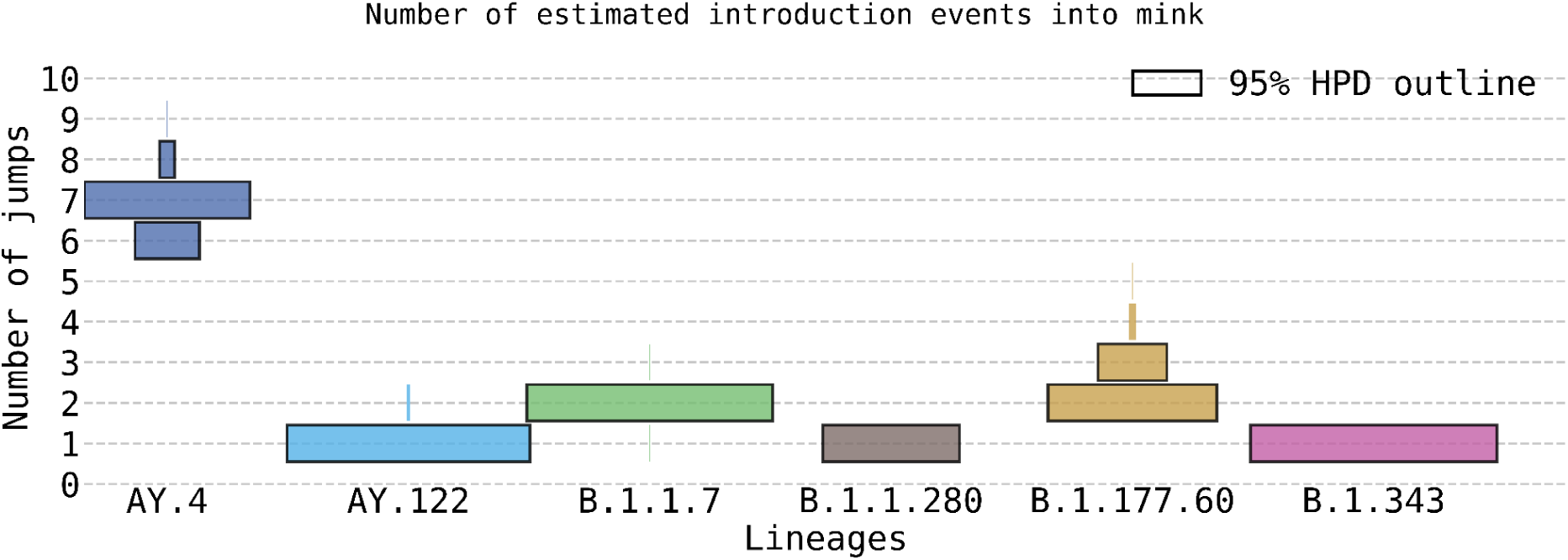
Posterior estimates of SARS-CoV-2 human-to-mink cross-species transmissions in Lithuania. The numbers of transitions (Y-axis) are based on Markov jumps from all generated MCMC output trees after burn-in removal, and coloured by lineage. The width of the coloured bar represents posterior probabilities of each number of introductions. The bars outlined in black encompass the 95% highest probability mass. AY.4 lineages exhibit the highest number of human-to-mink transmissions, with all other lineages having 1-3 detected anthropozoonotic jumps.

**Figure 5.**
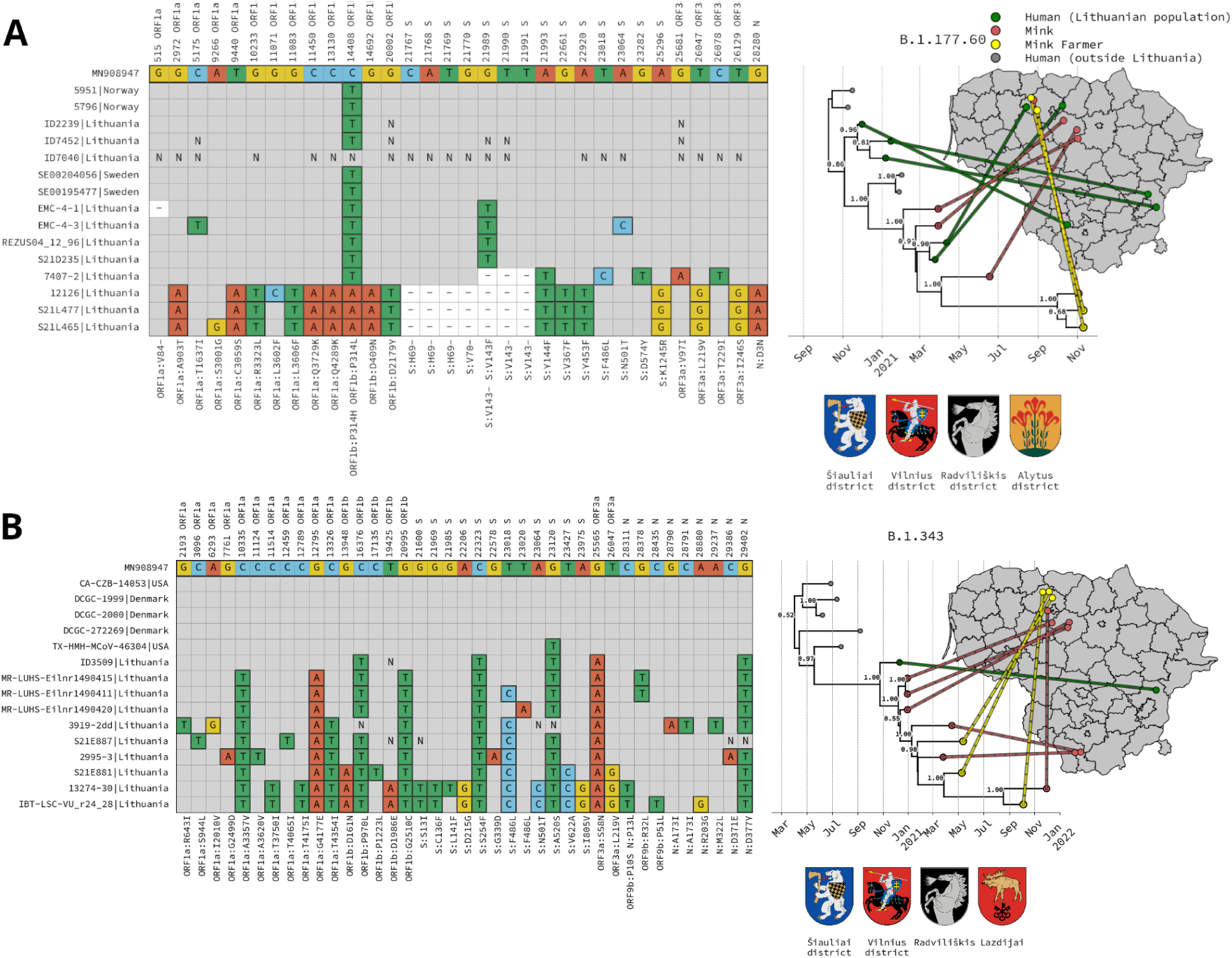
SNP alignments with phylogenetic trees. Starting from the left: A condensed alignment (described in methods) of polymorphic sites is displayed, keeping only the mutations causing amino acid changes or deletions in mink-derived SARS-CoV-2 genomes for lineages B.1.177.60 (A) and B.1.343 (B). Further to the right, subtrees containing the mink samples, extracted from MCC trees (with the closest contextual samples). Genomes in the alignment and tree are ordered the same. A map of Lithuania is shown on the right with lines connecting each tip to a random point within the municipality where the sample was collected. The coats of arms of municipalities where samples were collected are displayed below the tree.

#### B.1.177.60

Lineage B.1.177.60 circulated in the Lithuanian population from October 2020 to March 2021, and was a locally prevalent descendant of a common European lineage B.1.177 (Hodcroft et al., 2021). We estimate at least two spillover events from humans to mink between November 2020 and February 2021 (95% HPD: 2020 Nov 23 – 2021 Feb 19; Figure 3, Figure 4) and between December 2020 and November 2021 (95% HPD: 2020 Dec 04 – 2021 Nov 01; Figure 3, Figure 4). Although no human-to-human transmission of this lineage was observed in the general Lithuanian human population in late 2021, B.1.177.60 infections were identified in both mink and mink farm workers. Some sequences in this clade from human infections contain known mink-adaptive mutations (e.g., ORF3a: L219V (Tan et al., 2022)).

#### B.1.1.7

Lineage B.1.1.7 was prevalent in the Lithuanian population from January to June 2021, and we estimate at least two introductions of it from humans to mink between November 2020 and December 2021 (95% HPD: 2020 Nov 26 – 2021 Dec 04) and between January 2021 and August 2021 (95% HPD:2021 Jan 21 – 2021 Aug 23; Figure 3, Figure 4).

#### AY.4 and AY.122

Both lineages AY.4 and AY.122 circulated in the Lithuanian population from July 2021 to Jan 2022. We estimate at least seven (95% HPD: 2021 Jul 09 – 2021 Oct 04; 2021 Aug 07 – 2021 Nov 23; 2021 Aug 13 – 2021 Oct 27; 2021 Aug 15 – 2021 Nov 03; 2021 Sep 05 – 2021 Dec 04; 2021 Sep 09 – 2021 Nov 05; 2021 Oct 28 – 2022 May 16; (Figure 3, Figure 4) anthroponotic transmissions from humans to farmed mink of lineage AY.4 and one for AY.122 (95% HPD: 2021 Aug 31 – 2021 Oct 21; Figure 3, Figure 4). For AY.4, the 95% HPD regions, while overlapping, span from August 2021 to May 2022 with almost all of these jumps clustered around October 2021, at the time when AY.4 and descendant lineages were most common in humans. As for AY.122 lineage, the 95% highest posterior density region for the human to mink jump event is around September to October 2021, also coinciding with high numbers of SARS-CoV-2 infections in humans and peak farmed mink population within the year (Figure 3). We suspect that the volume of introductions of Delta-related lineages into mink is not inherently unique or reflects any changes in the underlying epidemiology of SARS-CoV-2 in either humans or mink but captures a period of time when Delta-like lineages were common in humans in Lithuania and mink were under intensive surveillance. As such, we believe that human-origin SARS-CoV-2 lineages in Lithuania probably spill over into farmed mink on a regular basis but remain undetected. Many such lineages in mink probably go extinct stochastically and the few that persist can only be caught during active veterinary surveillance which in this case was triggered in response to more successful mink-associated lineages (e.g. B.1.343 or B.1.177.60) jumping into and being discovered in humans.

### Mink-to-human transmissions

We identified around 4 independent SARS-CoV-2 mink-to-human transitions by inspecting MCC trees (Figure 5, Supplementary folder “MCC_Trees”): At least one for B.1.177.60 and most likely three for B.1.343 lineage. For B.1.177.60, two human samples (S21L477|Lithuania and S21L465|Lithuania) were sampled on the same day (2021-Nov-11) and it is impossible to determine if one of the infections was human-to-human, but they differ by one nucleotide (and one additional compatible but ambiguous nucleotide position) and are phylogenetically closely related to the 12126|Lithuania (EPI_ISL_10571267) sequence obtained from farmed mink (2021-Nov-04) (Figure 5). Human cases of B.1.343 in 2021, however, are not closely related, indicating the occurrence of multiple mink-to-human transmissions. Overall, the real number of mink-to-human jumps remains unclear as we have very limited sequence data from mink, especially close to their suspected transmission times. As can be seen in the AY.4 lineage group example (Figure 3; Figure 4; Supplementary Figures S1-S4) active veterinary surveillance readily detects multiple human-origin SARS-CoV-2 lineages in mink when these lineages are common in humans. However, once introduced into mink, these lineages rarely seem to establish themselves long-term, given that only a handful of lineages were detected in mink after their extinction in humans in Lithuania. Therefore, we expect a considerable number of mink infections with SARS-CoV-2 to have gone undetected, due to limited genomic surveillance efforts.

### Omicron lineages and further surveillance

Soon after the one pulse of active country-wide veterinary surveillance on mink farms for SARS-CoV-2, Omicron (B.1.1.529) and its descendants swept through the human population. In the absence of additional SARS-CoV-2 surveillance in mink after 2021, we do not have data to determine if Omicron-related lineages also entered mink populations and/or were able to establish and adapt to the mink populations.

## Discussion

Instances of SARS-CoV-2 introduction from humans into farmed mink (Jahid et al., 2024; Oreshkova et al., 2020), frequently prolonged and sustained circulation in mink (Domańska-Blicharz et al., 2023), and occasional anthropozoonotic jumps of SARS-CoV-2 from mink back into humans (Rabalski et al., 2022) have been documented throughout the COVID-19 pandemic. Here, using whole-genome sequencing data from routine Lithuanian SARS-CoV-2 genomic surveillance programme in humans, as well as a pulse of active veterinary surveillance in mink, we inferred the counts, timing and duration of such cross-species transmission events that were detected between 2021 and 2022 during the COVID-19 pandemic.

### Active and passive surveillance of farmed mink

The timing of active veterinary surveillance in mink (2021 November-December) led to a detection of 14 human-to-mink transmission events, with multiple recent transmissions for prevalent lineages at the time. As such, we suspect that the high number of introductions of specifically Delta lineages into mink (common in the Lithuanian population in late 2021) are representative of a trend where SARS-CoV-2 spillovers from humans into farmed mink in Lithuania happened continuously and in abundance but, as the B.1.343 and B.1.177.60 lineages demonstrate, only rarely established themselves and circulated for prolonged periods of time in mink. The few successful lineages continued circulating in mink undetected by passive veterinary surveillance until spillbacks into humans were detected by active human surveillance in Lithuania. These results are in agreement with previous research that found self-reporting-based passive veterinary surveillance as insufficient for detection of SARS-CoV-2 circulation in mink farms (Domańska-Blicharz et al., 2023; Žigaitė et al., 2023).

In addition, multiple other shortcomings in SARS-CoV-2 outbreak prevention in Lithuanian mink farms can be identified - farmers had no incentives to report outbreaks in mink farms (“Delfi Agro,” n.d.), decisive interventions (e.g. culling of affected farms) were never taken, active veterinary surveillance in mink farms was never routine and only implemented as a one-off intervention at the end of 2021 in response to spillback into humans, and little was done to communicate about the dangers of continued SARS-CoV-2 circulation in animals to the public. Furthermore, the European Commission published a document laying down rules for the monitoring and reporting of SARS-CoV-2 infections in animals which required active veterinary surveillance (“Implementing decision - 2021/788 - EN - EUR-Lex,” n.d.), while Lithuanian public institutions deemed that risk reduction measures implemented in mink farms were sufficient. This flawed implementation of veterinary surveillance gave a false impression about SARS-CoV-2 in Lithuanian mink farms which was severe and which we were able to uncover here. As such, and in the absence of ongoing active veterinary surveillance, the ban on mink farming, set to come into force in 2027 in Lithuania, seems to be the only viable solution for preventing SARS-CoV-2 circulation in mink.

In Lithuania, and many other countries, human and animal health are treated separately: institutions responsible for either rarely communicate or share data, and their responsibilities do not directly overlap; therefore, coordinating a response in cases of zoonosis or conducting epidemiological studies for pathogens infecting both human and animal hosts, as shown by our study, presents an immense challenge.

The One Health approach, proposed and promoted by the World Health Organisation, is crucial when addressing diseases that pose threats to both humans and animals (Mackenzie and Jeggo, 2019; “One Health,” n.d.). Notably, there have been success stories in combining both animal and human health: rabies human cases in Latin America were drastically reduced by vaccinating both humans and canines (Freire de Carvalho et al., 2018). In Malaysia, the Nipah virus outbreak was controlled only as it was correctly identified, found in pigs, and pig culling was introduced (Looi and Chua, 2007). *M.ulcerans* clusters in southeastern Australia were shown to be overlapping among humans and mosquitoes, prompting mosquito control as a viable measure against *M.ulcerans* infections (Mee et al., 2024).

### Situation in other countries

Many developed nations have reported SARS-CoV-2 outbreaks in mink farms - including Netherlands, Spain, Denmark, United States, Sweden, Italy, France, Greece, Lithuania, Canada, Poland, and Latvia (Jahid et al., 2024; Oreshkova et al., 2020). In some cases there were secondary spillover events back to humans, for example in the Netherlands where it was demonstrated that some mink farmers were infected with a mink-associated lineage instead of what was prevalent in the local human population (Lu et al., 2021) or in Denmark, where a mink associated lineage circulated in local human population (Fenollar et al., 2021; Rasmussen et al., 2024). In Poland, it was demonstrated that SARS-CoV-2 outbreaks in mink can last for months, by detection of cryptic lineages in mink farms which were not found in humans (Domańska-Blicharz et al., 2023). In both Denmark and the Netherlands, mass culling was initiated, preventing future backspill events, while the outcome of the situation in Poland remains unknown. We suspect that human-to-mink transmission was common in Lithuania, along with subsequent mink-to-human transmission. However, it is highly likely that these transmission events are not always detected or reported; therefore, the number of outbreaks and their severity in mink farms remains underestimated. A glance at GISAID data from Latvia, a neighboring country, suggests a similar situation. Lineage B.1.177.60 was detected in mink from April to November 2021, while human samples of this lineage were collected from September 2020 to July 2021. One human B.1.177.60 sample from October 2021 (uploaded by the Institute of Food Safety, Animal Health, and Environment of Latvia), a lineage almost certainly extinct in humans by that point, strongly suggests the individual may have worked on a mink farm, though this information is not available to us.

### Surveillance gaps in Lithuania

Our analysis revealed multiple likely human-to-mink SARS-CoV-2 jumps during the COVID-19 pandemic, some of which circulated undetected within mink populations for extended periods and resulted in anthropozoonotic spillovers back to humans. Essentially, our findings highlight critical gaps in the passive surveillance (especially genomic) system, hindered by the absence of incentives for self-reporting and insufficient communication between human and animal health institutions. Ultimately, the lack of decisive action from institutions responsible for animal health can pose a serious threat to human health and addressing these issues is crucial to improve preparedness and response during zoonotic outbreaks in future pandemics.

## Data Availability

GISAID acknowledgment table, list of accessions, relevant data from Lithuania, analysis input and output files, and scripts used to process and visualise data are available at Zenodo, DOI: 10.5281/zenodo.15858336.

## Data availability

Main and supplementary figures, BEAST XML (with sequences removed, per GISAID’s user agreement), trees (posterior and MCC), and log files, data on mink and human SARS-CoV-2 case data, GISAID acknowledgment tables and accessions used, as well as code to analyse and visualise data are available on Zenodo: DOI: 10.5281/zenodo.15858336.

## Acknowledgments

GD acknowledges the support of EMBO installation grant EMBO-IG-5305-2023.

## Supplementary materials

Data supporting the findings of this study are available within the article and its supplementary files. Supplementary files are through Zenodo (DOI: 10.5281/zenodo.15858336).

### Additional alignments

**Figure S1.**
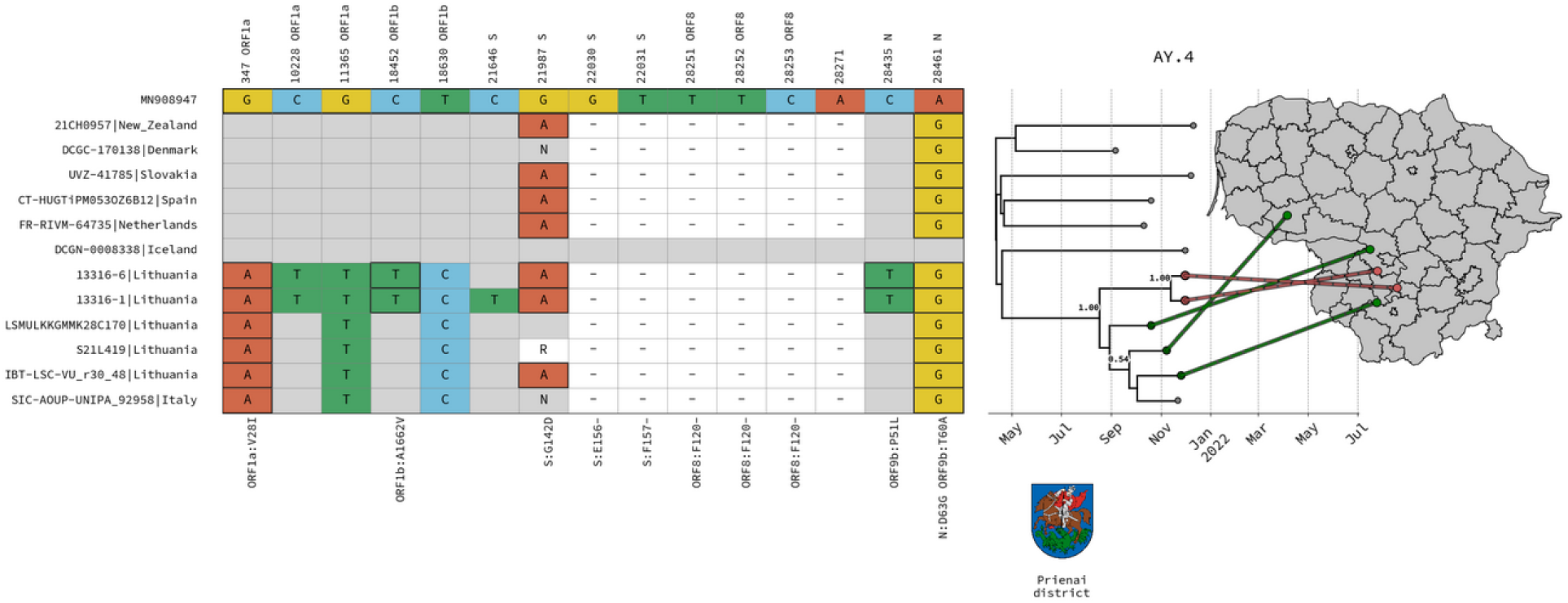
SNP alignments with phylogenetic trees. Starting from the left: A condensed alignment of polymorphic sites is displayed, keeping only the mutations in mink samples for lineage AY.4 (One of multiple alignments). Further to the right, subtrees containing the mink samples, extracted from MCC trees (with the closest contextual samples). A map of Lithuania with lines from the leaf in a tree to a point on a map, representing the municipality of origin of a given sample (points are randomly positioned within the area of the municipality). The coats of arms of municipalities where mink samples were collected are displayed below the tree.

**Figure S2.**
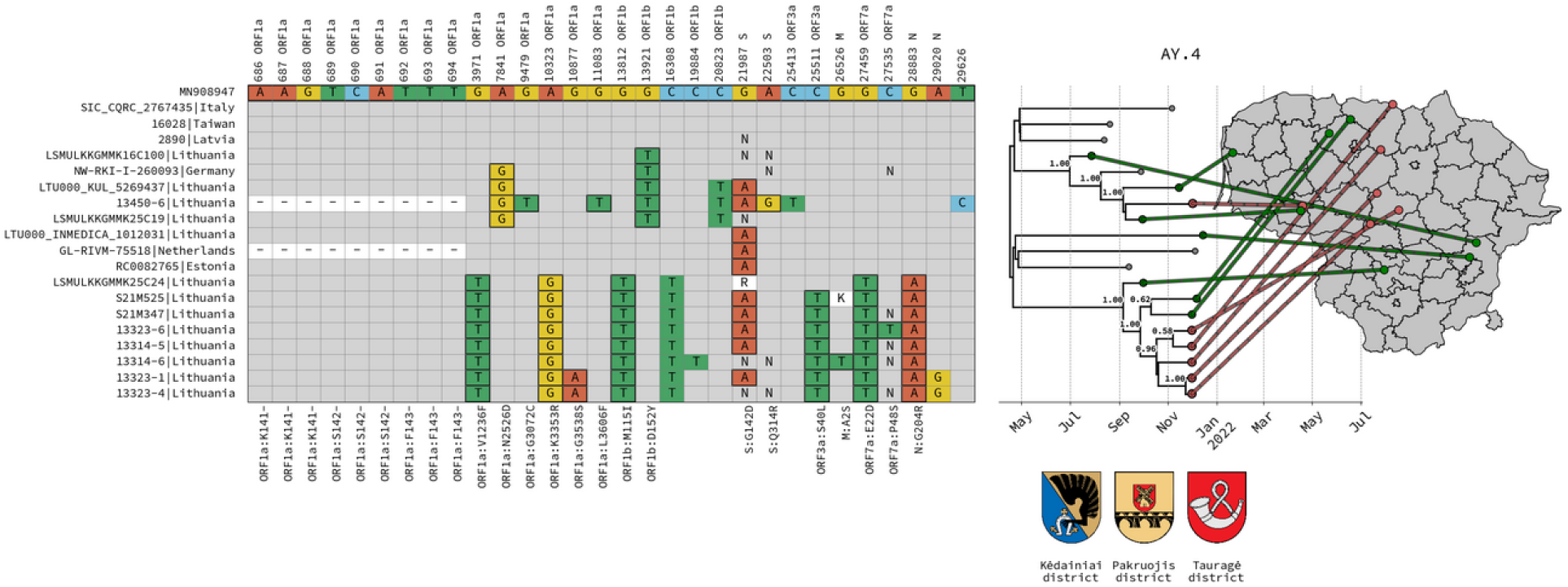
SNP alignments with phylogenetic trees. Starting from the left: A condensed alignment of polymorphic sites is displayed, keeping only the mutations in mink samples for lineage AY.4 (One of multiple alignments). Further to the right, subtrees containing the mink samples, extracted from MCC trees (with the closest contextual samples). Additionally, a map of Lithuania with lines from the leaf in a tree to a point on a map, representing the municipality of origin of a given sample (points are randomly positioned within the area of the municipality if municipality data is available). The coats of arms of municipalities where mink samples were collected are displayed below the tree.

**Figure S3.**
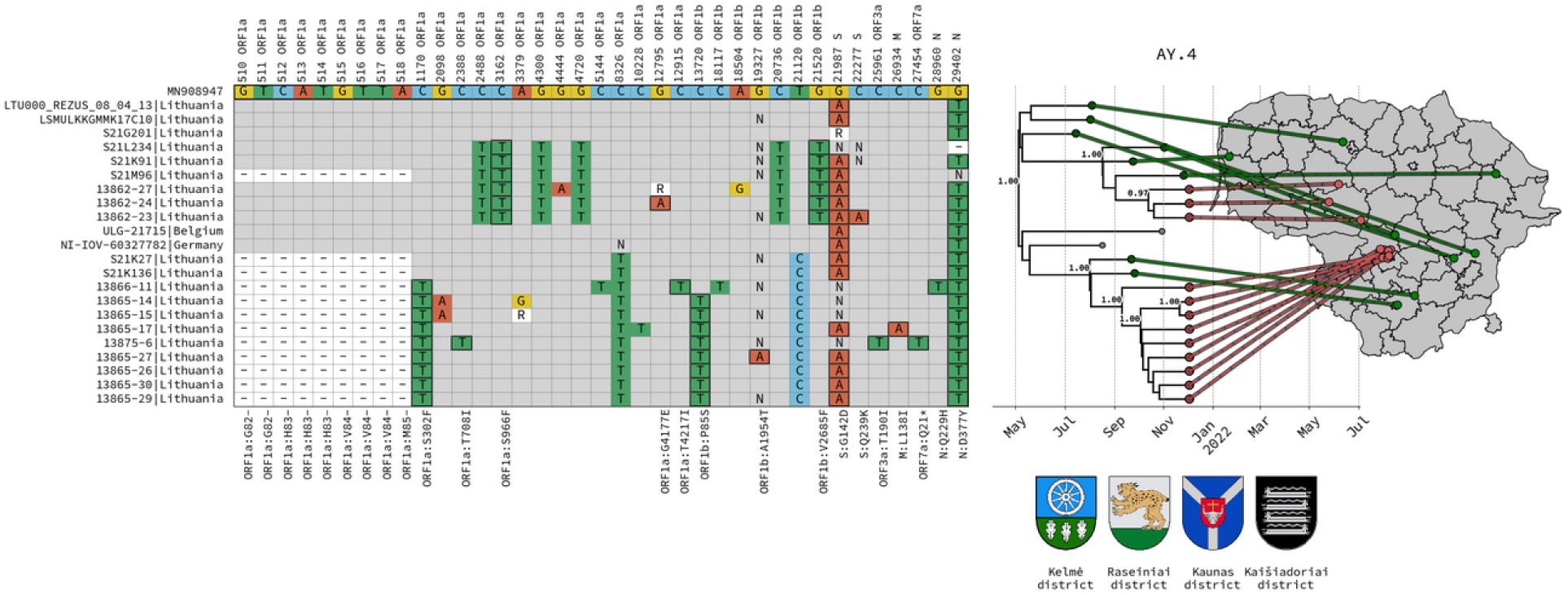
SNP alignments with phylogenetic trees. Starting from the left: A condensed alignment of polymorphic sites is displayed, keeping only the mutations in mink samples for lineage AY.4 (One of multiple alignments). Further to the right, subtrees containing the mink samples, extracted from MCC trees (with the closest contextual samples). Additionally, a map of Lithuania with lines from the leaf in a tree to a point on a map, representing the municipality of origin of a given sample (points are randomly positioned within the area of the municipality). The coats of arms of municipalities where mink samples were collected are displayed below the tree.

**Figure S4.**
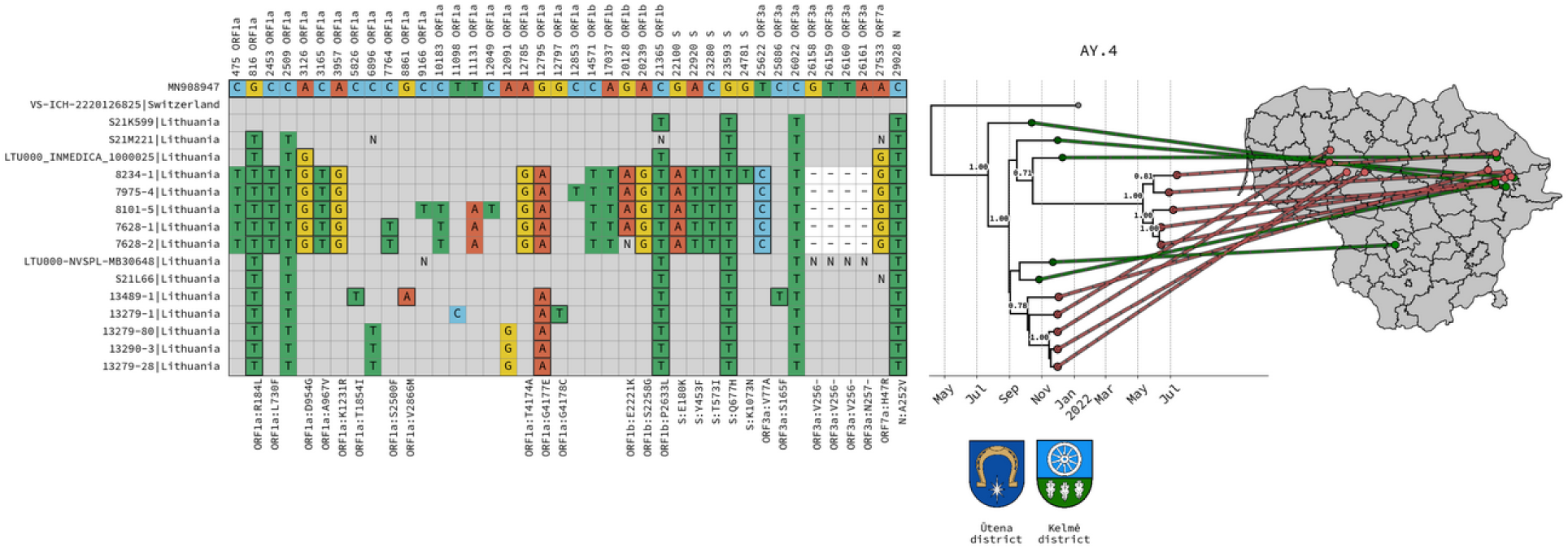
SNP alignments with phylogenetic trees. Starting from the left: A condensed alignment of polymorphic sites is displayed, keeping only the mutations in mink samples for lineage AY.4 (One of multiple alignments). Further to the right, subtrees containing the mink samples, extracted from MCC trees (with the closest contextual samples). Additionally, a map of Lithuania with lines from the leaf in a tree to a point on a map, representing the municipality of origin of a given sample (points are randomly positioned within the area of the municipality). The coats of arms of municipalities where mink samples were collected are displayed below the tree.

**Figure S5.**
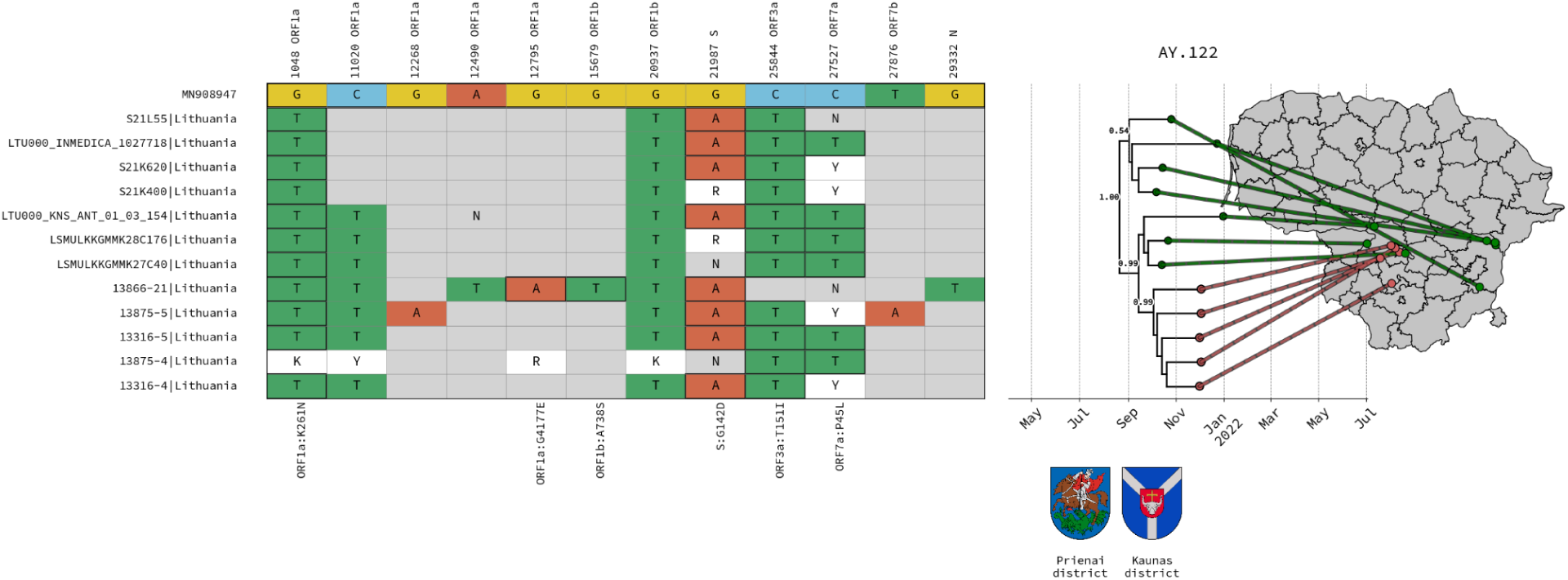
SNP alignments with phylogenetic trees. Starting from the left: A condensed alignment of polymorphic sites is displayed, keeping only the mutations in mink samples for lineage AY.122. Further to the right, subtrees containing the mink samples, extracted from MCC trees (with the closest contextual samples). Additionally, a map of Lithuania with lines from the leaf in a tree to a point on a map, representing the municipality of origin of a given sample (points are randomly positioned within the area of the municipality). The coats of arms of municipalities where mink samples were collected are displayed below the tree.

**Figure S6.**
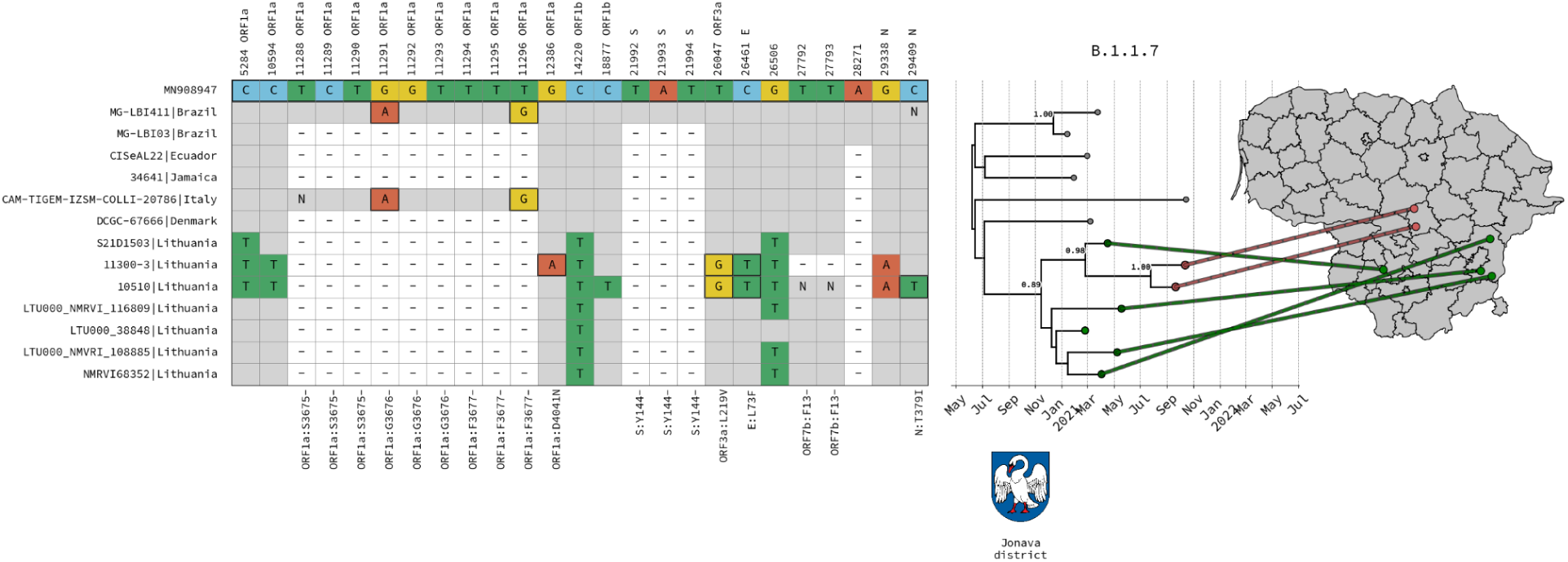
SNP alignments with phylogenetic trees. Starting from the left: A condensed alignment of polymorphic sites is displayed, keeping only the mutations in mink samples for lineage B.1.1.7. (One of two alignments). Further to the right, subtrees containing the mink samples, extracted from MCC trees (with the closest contextual samples). Additionally, a map of Lithuania with lines from the leaf in a tree to a point on a map, representing the municipality of origin of a given sample (points are randomly positioned within the area of the municipality if municipality data is available). The coats of arms of municipalities where mink samples were collected are displayed below the tree.

**Figure S7.**
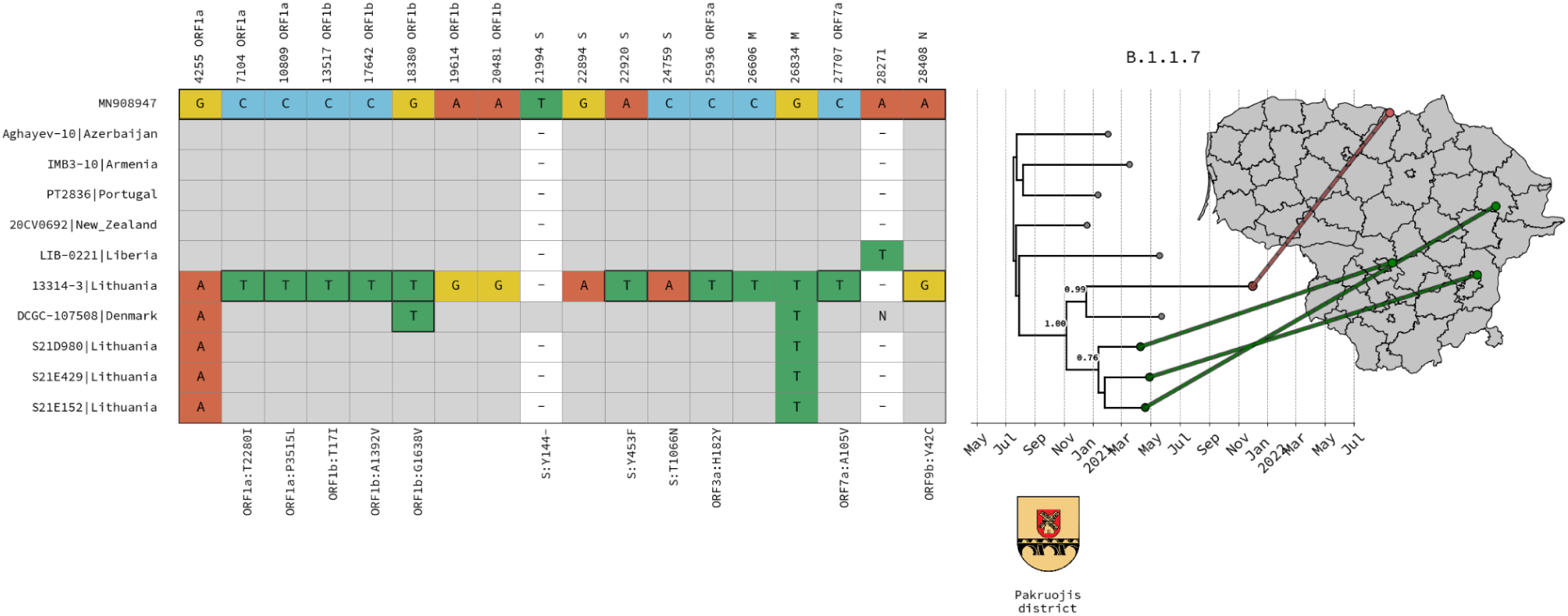
SNP alignments with phylogenetic trees. Starting from the left: A condensed alignment of polymorphic sites is displayed, keeping only the mutations in mink samples for lineage B.1.1.7. (One of two alignments). Further to the right, subtrees containing the mink samples, extracted from MCC trees (with the closest contextual samples). Additionally, a map of Lithuania with lines from the leaf in a tree to a point on a map, representing the municipality of origin of a given sample (points are randomly positioned within the area of the municipality). The coats of arms of municipalities where mink samples were collected are displayed below the tree.

**Figure S8.**
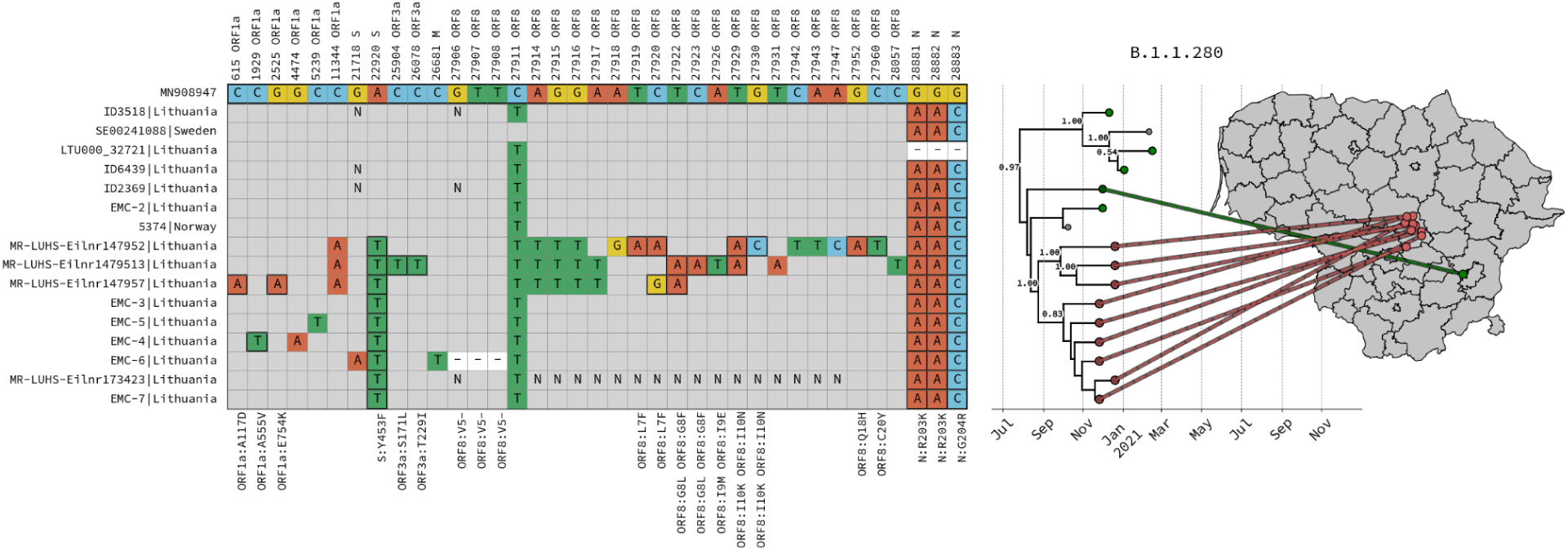
SNP alignments with phylogenetic trees. Starting from the left: A condensed alignment of polymorphic sites is displayed, keeping only the mutations in mink samples for lineage B.1.1.280. Further to the right, subtrees containing the mink samples, extracted from MCC trees (with the closest contextual samples). Additionally, a map of Lithuania with lines from the leaf in a tree to a point on a map, representing the municipality of origin of a given sample (points are randomly positioned within the area of the municipality). The coats of arms of municipalities where mink samples were collected are displayed below the tree.

**Figure S9.**
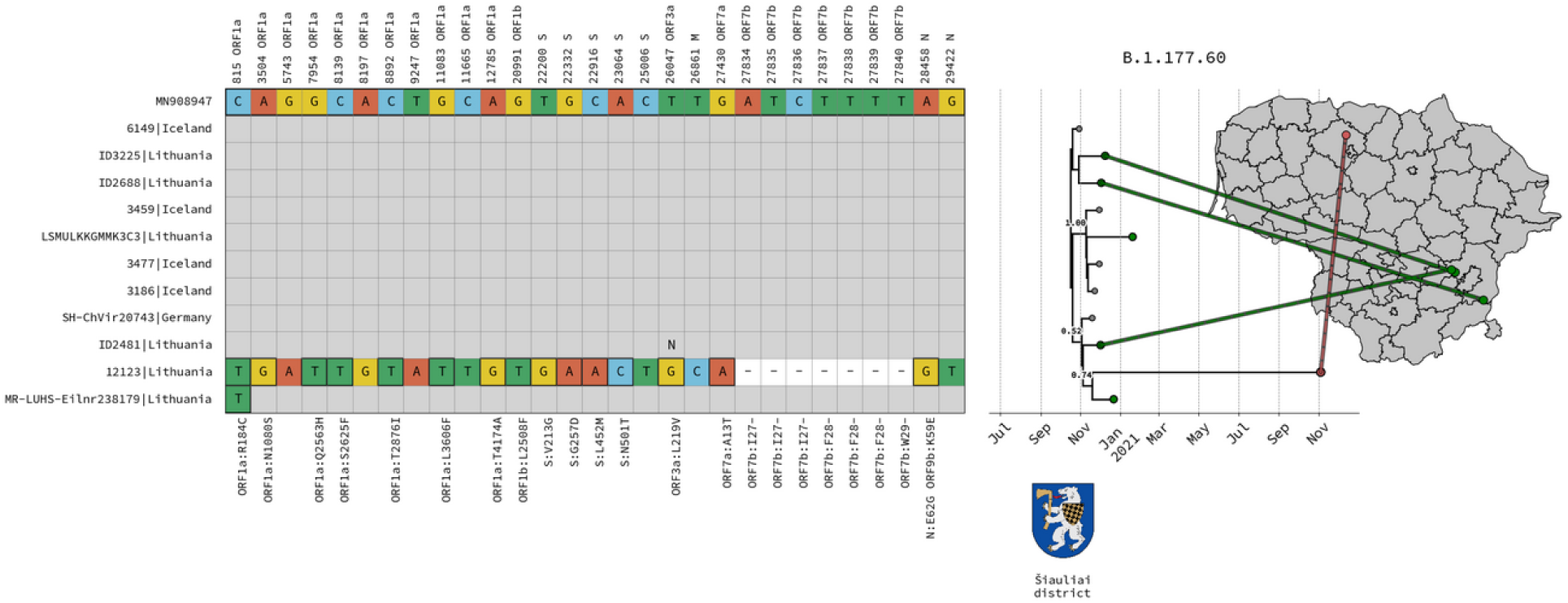
SNP alignments with phylogenetic trees. Starting from the left: A condensed alignment of polymorphic sites is displayed, keeping only the mutations in mink samples for lineage B.1.177.60 secondary transitions. Further to the right, subtrees containing the mink samples, extracted from MCC trees (with the closest contextual samples). Additionally, a map of Lithuania with lines from the leaf in a tree to a point on a map, representing the municipality of origin of a given sample (points are randomly positioned within the area of the municipality if municipality data is available). The coats of arms of municipalities where mink samples were collected are displayed below the tree.

### Biosecurity requirements in Lithuanian mink farms

Biosecurity requirements issued by the SFVS have been compulsory on mink farms in the country since 2015 (“B1-432,” n.d.). The main requirements were: 1) fencing of farms to keep out wildlife and unauthorized persons (likely ineffective (Sikkema et al., 2022)), 2) staff must wear only mink dedicated work clothes and footwear, 3) use of disinfection barriers, handwashing and hand disinfection equipment, 4) regular rodent and pest control, in addition to disinfection of vehicles while entering and exiting the premises through changing rooms with showers were recommended. Furthermore, the responsible person on a mink farm has to be informed if a visitor has visited other mink premises in the last 48 hours, has been on a fox or raccoon dog hunt, has been in contact with animal by-products, in which case the person responsible for the mink farm will decide whether such a visitor may be admitted to the holding. However, it is unclear how compliance with these requirements is regulated.

### Description of statistical data

#### Kernel Density Estimation (KDE)

We used Python script to calculate KDE from posterior distributions of BEAST data regarding human-to-mink transition dates for each lineage. We employed Gaussian KDE from the scipy package (scipy.stats.gaussian_kde) with bandwidth modified by 0.8 (kde.set_bandwidth(bw_method=kde.factor * 0.8). We set the evaluation at a thousand points for each case (x_kde = np.linspace(min(data), max(data), 1000)) and calculating each value on Y-axis (y_kde = kde(x_kde)) with further normalization for more even visualization, based on total area under the curve (dx = x_kde[1] - x_kde[0]; y_kde_normalized = y_kde / (np.sum(y_kde) * dx * 2000))

#### Highest Density Region (HDR) estimation

We used HDR calculation regarding transition from human to mink event number and dates for each lineage. We used a custom Python script, by first using Gaussian KDE (scipy.stats.gaussian_kde) to count and bin x-values spanning the sample range, by creating a probability density function. HDR was defined as a union of shortest intervals that together contain 95%.

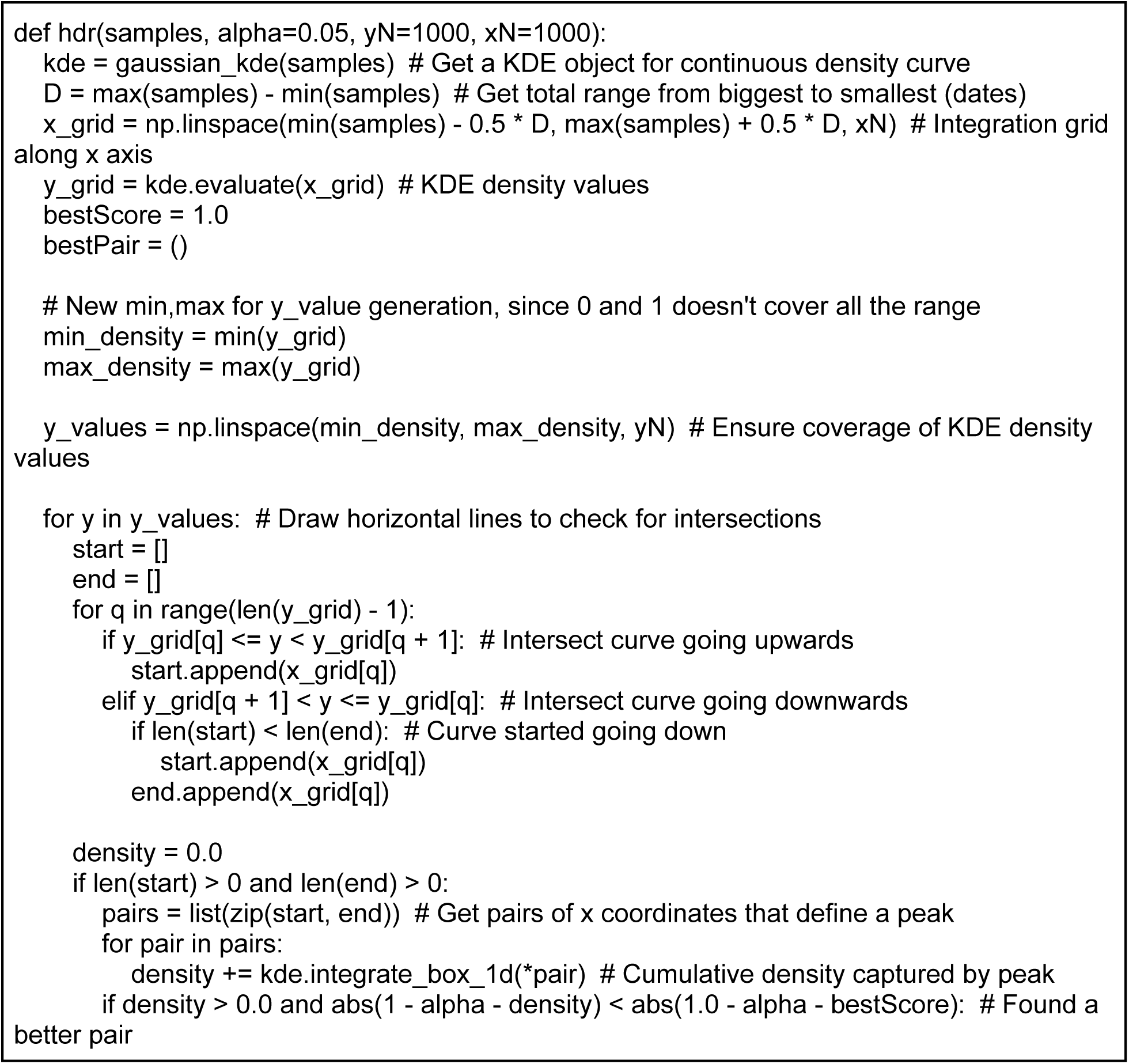
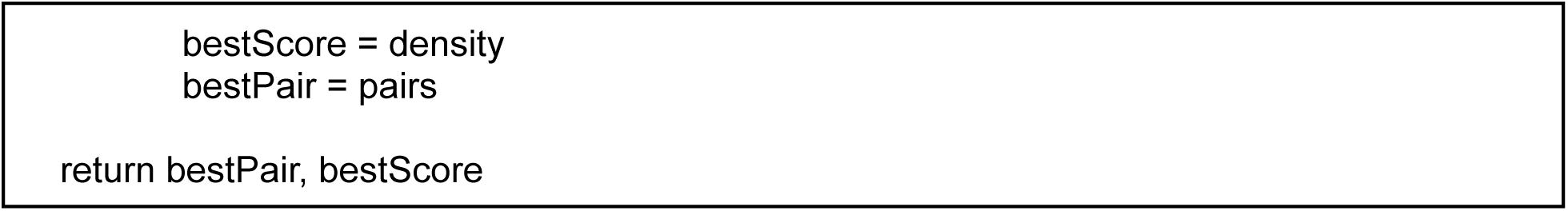

